# Multi-ethnic Investigation of Risk and Immune Determinants of COVID-19 Outcomes

**DOI:** 10.1101/2021.10.29.21265687

**Authors:** Tomi Jun, Divij Mathew, Navya Sharma, Sharon Nirenberg, Hsin-Hui Huang, Patricia Kovatch, E. John Wherry, Kuan-lin Huang

**Author notes:** Corresponding Author: Kuan-lin Huang, Ph.D., Department of Genetics and Genomic Sciences, Icahn School of Medicine at Mount Sinai, New York, NY 10029.

## Abstract

**Objectives:** To compare risk factors for COVID-19 mortality among hospitalized Hispanic, Non-Hispanic Black, and White patients.

**Design:** Retrospecitve cohort study

**Setting:** Five hosptials within a single academic health system

**Participants:** 3,086 adult patients with self-reported race/ethnicity information presenting to the emergency department and hospitalized with COVID-19 up to April 13, 2020.

**Main outcome measures:** In-hospital mortality

**Results:** While older age (multivariable OR 1.06, 95% CI 1.05-1.07) and baseline hypoxia (multivariable OR 2.71, 95% CI 2.17-3.36) were associated with increased mortality overall and across all races/ethnicities, Non-Hispanic Black (median age 67, IQR 58-76) and Hispanic (median age 63, IQR 50-74) patients were younger and had different comorbidity profiles compared to Non-Hispanic White patients (median age 73, IQR 62-84; p<0.05 for both comparisons). Among inflammatory markers associated with COVID-19 mortality, there was a significant interaction between the Non-Hispanic Black population and interleukin-1-beta (interaction p-value 0.04).

**Conclusions:** This analysis of a multi-ethnic cohort highlights the need for inclusion and consideration of diverse popualtions in ongoing COVID-19 trials targeting inflammatory cytokines.

## Introduction

Reports from the United States, the United Kingdom, and Brazil have highlighted racial disparities in the COVID-19 pandemic.^1–4^ National studies from the UK and Brazil have found race to be an independent predictor of death.^3,4^ In the United States, Black and Hispanic individuals have disproportionately high rates of infection, hospitalization, and mortality.^1,2,5,6^ These disparities have been attributed to greater representation of Black and Hispanic persons in essential services, and a higher burden of comorbidities in minority communities, among others.

While Black and Hispanic individuals in the US have been disproportionately affected by the pandemic, the majority of published studies investigating COVID-19 mortality risk factors have been in cohorts of individuals with predominantly European or Asian ancestry.^7–11^ Few US studies have directly examined mortality risk factors and their effect sizes in Black or Hispanic compared to White individuals.^1,12^ Rigorous analysis to establish risk factors and molecular predictors for each population is urgently needed.

We sought to identify race/ethnic-specific clinical and immune factors of mortality using a diverse cohort of White, Black, and Hispanic COVID-19 patients admitted to a single health system in New York. In addition to baseline characterization, we conducted stratified and interaction term analyses to identify risk and immune factors that may affect outcomes of each patient population. The systematic analyses revealed population-specific effects of multiple risk factors that were previously unknown, highlighting the importance of including diverse patient populations and tailored consideration in precision medicine for COVID-19.

## Results

### Study population

There were 4,997 adult patients with COVID-19-related emergency department (ED) visits on or before April 13^th^, 2020, of whom 3,086 (61.8%) were hospitalized. Hospitalization rates were significantly lower for Non-Hispanic (NH) Black patients compared to other groups (NH Black 56.5%; NH White 63.4%; Hispanic 64.3%, Asian 62.6%; Other 63.6%; p<0.001). Hospitalized patients were significantly older (median age 66 vs. 50, p<0.001) and were more likely to have comorbidities such as hypertension (35.5% vs. 13.5%, p<0.001), diabetes (24% vs. 7.8%, p<0.001), and chronic kidney disease (11.9% vs. 3.1%, p<0.001) (**Supplemental Table 1**). The clinical characteristics of non-hospitalized patients are summarized by race/ethnicity in **Supplemental Table 2**.

The hospitalized cohort included 3,086 adult patients. We excluded 78 patients with missing race or ethnicity data, 144 Asian patients, and 458 patients with other or unspecified race/ethnicity from the comparative/stratified analyses based on power considerations (**Figure 1**). The remaining 2,406 patients were 37.1% Hispanic, 34.3% Non-Hispanic (NH) Black, and 28.6% NH White.

**Figure 1:**
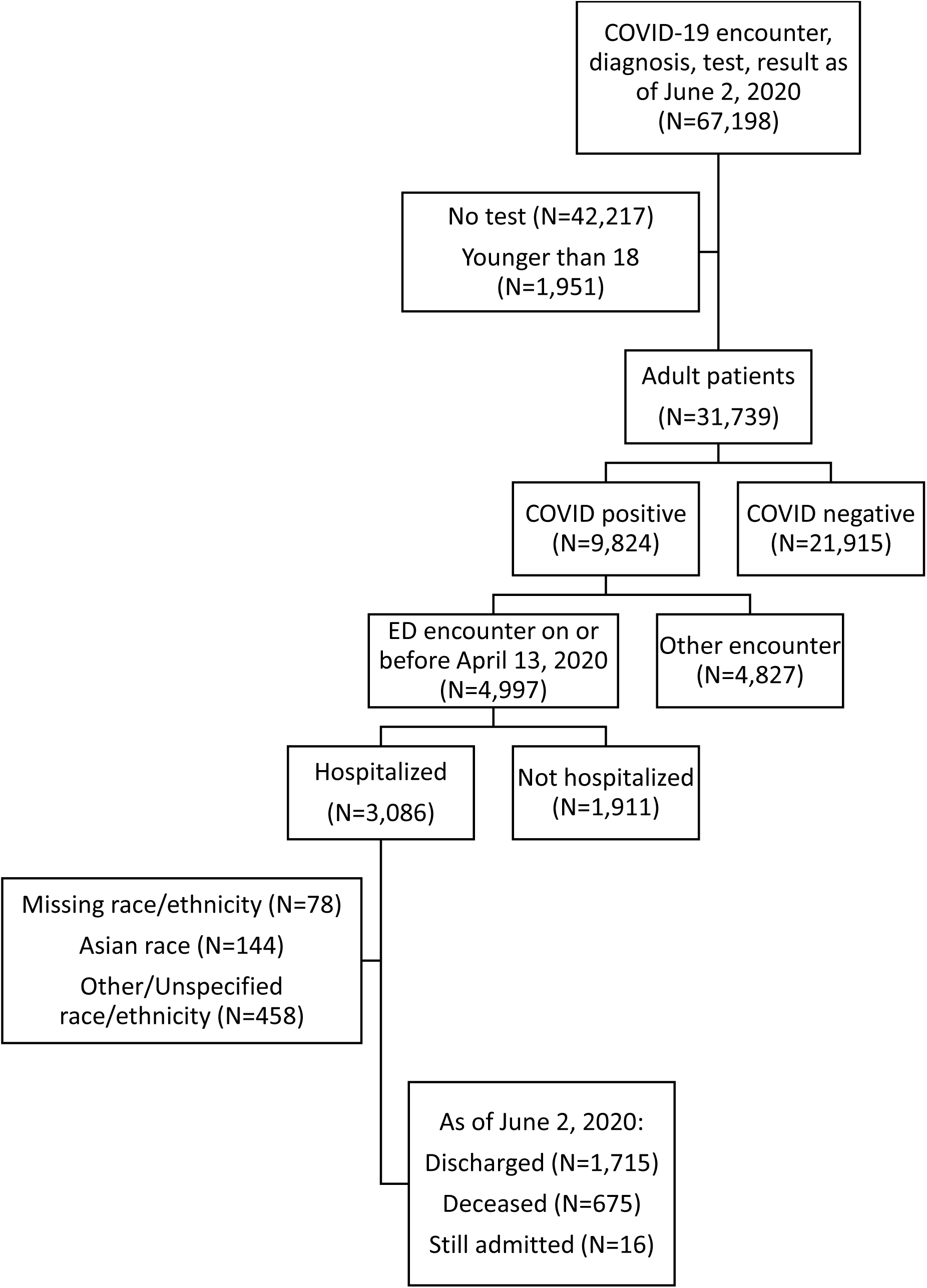
Flow diagram of included patients

Compared to NH White patients, NH Black and Hispanic patients were younger (median age 67 and 63 vs. 73, p<0.001 for both) (**Table 1**). NH Black patients were more likely to have hypertension (40.6% vs 31.8%, p<0.001), diabetes (26.8% vs. 17.4%, p<0.001), and chronic kidney disease (16.1% vs. 8%, p<0.001) than NH White patients. Hispanic patients were more likely to have diabetes (27.4% vs. 17.4%, p<0.001), chronic kidney disease (13% vs. 8%, p=0.001), and chronic liver disease (4.3% vs. 1.6%, p=0.003), compared to NH White patients. NH White patients were more likely to have coronary artery disease (17.1% vs. 12.2% and 11.3%, p=0.008 vs. Black, p=0.001 vs. Hispanic) and atrial fibrillation (12.3% vs. 5% and 4.5%, p<0.001 for both) than NH Black or Hispanic patients.

**Table 1:**
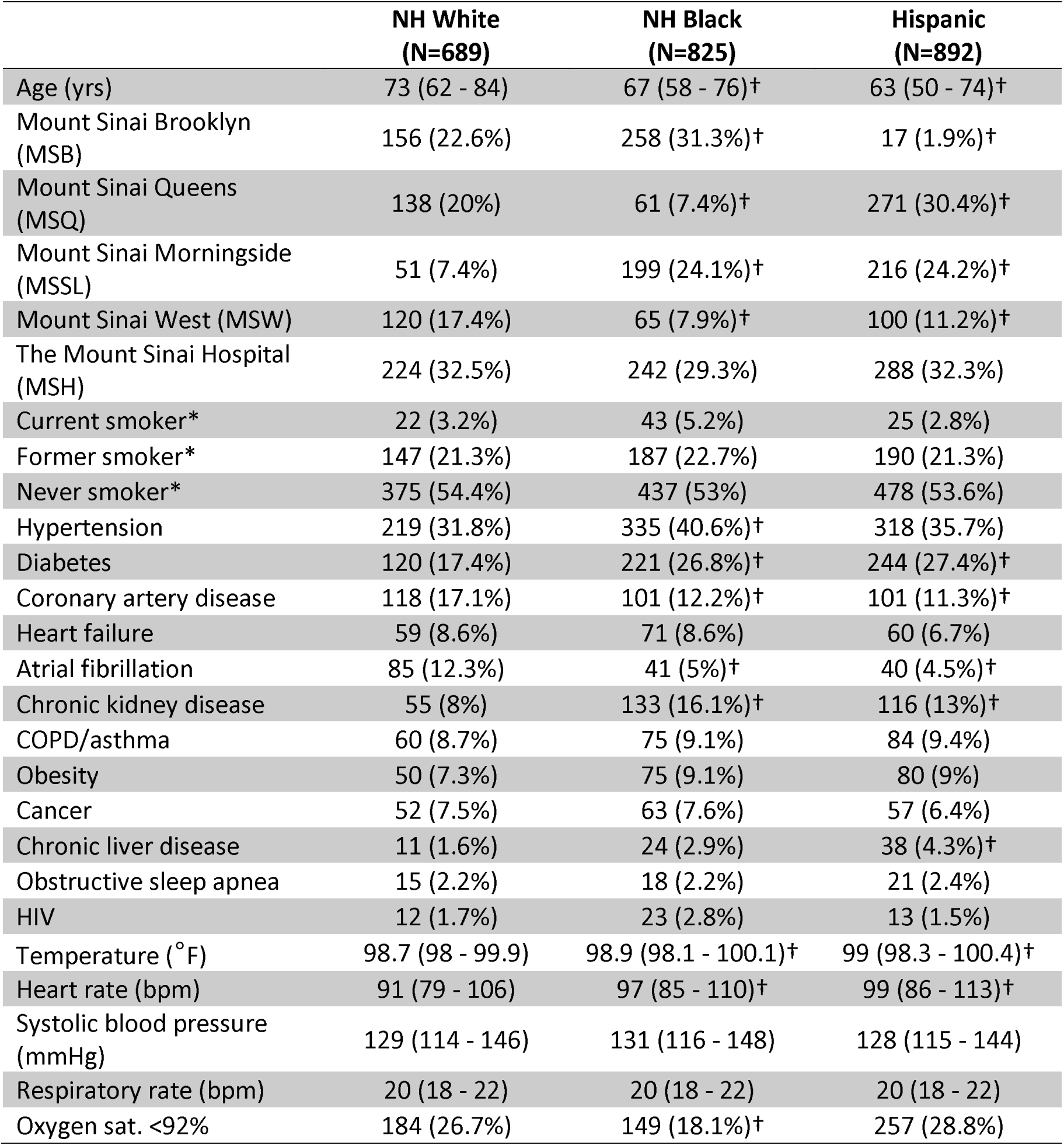
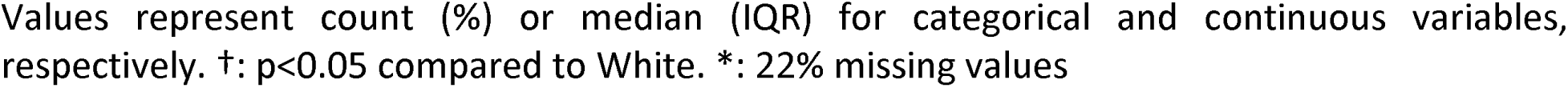
Baseline demographic and clinical characteristics, by race/ethnicity

|  | **NH White**<br>**(N=689)** | **NH Black**<br>**(N=825)** | **Hispanic**<br>**(N=892)** |
| --- | --- | --- | --- |
| Age (yrs) | 73 (62 - 84) | 67 (58 - 76)† | 63 (50 - 74)† |
| Mount Sinai Brooklyn (MSB) | 156 (22.6%) | 258 (31.3%)† | 17 (1.9%)† |
| Mount Sinai Queens (MSQ) | 138 (20%) | 61 (7.4%)† | 271 (30.4%)† |
| Mount Sinai Morningside (MSSL) | 51 (7.4%) | 199 (24.1%)† | 216 (24.2%)† |
| Mount Sinai West (MSW) | 120 (17.4%) | 65 (7.9%)† | 100 (11.2%)† |
| The Mount Sinai Hospital (MSH) | 224 (32.5%) | 242 (29.3%) | 288 (32.3%) |
| Current smoker* | 22 (3.2%) | 43 (5.2%) | 25 (2.8%) |
| Former smoker* | 147 (21.3%) | 187 (22.7%) | 190 (21.3%) |
| Never smoker* | 375 (54.4%) | 437 (53%) | 478 (53.6%) |
| Hypertension | 219 (31.8%) | 335 (40.6%)† | 318 (35.7%) |
| Diabetes | 120 (17.4%) | 221 (26.8%)† | 244 (27.4%)† |
| Coronary artery disease | 118 (17.1%) | 101 (12.2%)† | 101 (11.3%)† |
| Heart failure | 59 (8.6%) | 71 (8.6%) | 60 (6.7%) |
| Atrial fibrillation | 85 (12.3%) | 41 (5%)† | 40 (4.5%)† |
| Chronic kidney disease | 55 (8%) | 133 (16.1%)† | 116 (13%)† |
| COPD/asthma | 60 (8.7%) | 75 (9.1%) | 84 (9.4%) |
| Obesity | 50 (7.3%) | 75 (9.1%) | 80 (9%) |
| Cancer | 52 (7.5%) | 63 (7.6%) | 57 (6.4%) |
| Chronic liver disease | 11 (1.6%) | 24 (2.9%) | 38 (4.3%)† |
| Obstructive sleep apnea | 15 (2.2%) | 18 (2.2%) | 21 (2.4%) |
| HIV | 12 (1.7%) | 23 (2.8%) | 13 (1.5%) |
| Temperature (°F) | 98.7 (98 - 99.9) | 98.9 (98.1 - 100.1)† | 99 (98.3 - 100.4)† |
| Heart rate (bpm) | 91 (79 - 106) | 97 (85 - 110)† | 99 (86 - 113)† |
| Systolic blood pressure (mmHg) | 129 (114 - 146) | 131 (116 - 148) | 128 (115 - 144) |
| Respiratory rate (bpm) | 20 (18 - 22) | 20 (18 - 22) | 20 (18 - 22) |
| Oxygen sat. <92% | 184 (26.7%) | 149 (18.1%)† | 257 (28.8%) |

Values represent count (%) or median (IQR) for categorical and continuous variables, respectively. †: $p < 0.05$ compared to White. \*: 22% missing values

### Population-specific clinical factors associated with death

Unadjusted mortality rates were lower among NH Black (27.5% vs. 34.1%, p=0.006) and Hispanic (23.9% vs. 34.1%, p<0.001) patients compared to NH White patients. Rates of intensive care were not significantly different between NH Black and NH White (19.8% vs 21.5%, p=0.44) or Hispanic and NH White (23.4% vs 21.5%, p=0.36) patients.

We first evaluated the association of demographic, clinical, and laboratory variables with in-hospital mortality (**Table 2, Supplemental Table 4 & 5**). In a univariate analysis, NH Black (OR 0.73, 95% CI 0.59-0.91) and Hispanic (OR 0.61, 95% CI 0.49-0.76) populations were associated with lower mortality compared to NH White. However, race/ethnicity was not an independent predictor of mortality after adjusting for age, sex, comorbidities, and baseline hypoxia (oxygen saturation <92% at the first measurement of the clinical encounter) in this cohort (Black HR 1.03, 95% CI 0.80-1.32; Hispanic HR 0.94, 95% CI 0.73-1.21) (**Figure 2A**). Our finding is consistent with several cohort studies in the US,^1,13,14^ although UK and Brazil studies have reported race as an independent predictor of mortality,^3,4^ possibly due to population differences.

**Table 2:** Multivariable model predicting in-hospital mortality, stratified by race/ethnicity

| Variable | NH White OR<br>(95% CI) | NH Black OR<br>(95% CI) | Hispanic OR<br>(95% CI) | All patients OR<br>(95% CI) |
| --- | --- | --- | --- | --- |
| Male | 1.51 (1.03-2.22) | 1.55 (1.09-2.19) | 1.04 (0.73-1.49) | 1.29 (1.05-1.58) |
| Age (yrs) | 1.07 (1.05-1.09) | 1.06 (1.04-1.07) | 1.05 (1.04-1.07) | 1.06 (1.05-1.07) |
| Race: NH White | Reference | Reference | Reference | Reference |
| Race: NH Black | NA | NA | NA | 1.03 (0.80-1.32) |
| Race: Hispanic | NA | NA | NA | 0.94 (0.73-1.21) |
| Manhattan facility | 0.45 (0.31-0.64) | 0.53 (0.37-0.75) | 0.62 (0.43-0.89) | 0.53 (0.43-0.65) |
| Hypertension | 0.86 (0.56-1.33) | 0.69 (0.45-1.06) | 0.78 (0.51-1.2) | 0.77 (0.61-0.98) |
| Diabetes | 0.99 (0.61-1.63) | 1.67 (1.09-2.58) | 1.21 (0.79-1.83) | 1.26 (0.98-1.63) |
| Coronary artery disease | 0.98 (0.58-1.65) | 1.14 (0.67-1.93) | 0.10 (0.57-1.73) | 1.03 (0.76-1.39) |
| Heart failure | 0.94 (0.48-1.85) | 1.11 (0.6-2.06) | 1.14 (0.55-2.36) | 1.05 (0.72-1.54) |
| Atrial fibrillation | 1.15 (0.66-2.02) | 1.27 (0.62-2.59) | 1.99 (0.97-4.11) | 1.38 (0.95-2.00) |
| Chronic kidney disease | 2.25 (1.18-4.28) | 1.71 (1.06-2.77) | 1.38 (0.80-2.38) | 1.70 (1.25-2.31) |
| COPD/asthma | 0.9 (0.48-1.68) | 0.80 (0.42-1.54) | 0.58 (0.32-1.06) | 0.75 (0.52-1.07) |
| Obesity | 0.72 (0.33-1.60) | 1.33 (0.69-2.58) | 1.59 (0.86-2.91) | 1.25 (0.86-1.83) |
| Cancer | 0.95 (0.49-1.85) | 1.15 (0.61-2.16) | 1.65 (0.88-3.07) | 1.25 (0.86-1.8) |
| Oxygen sat. <92% | 2.377 (1.6-3.54) | 2.51 (1.69-3.75) | 3.22 (2.24-4.61) | 2.71 (2.17-3.36) |

**Figure 2:**
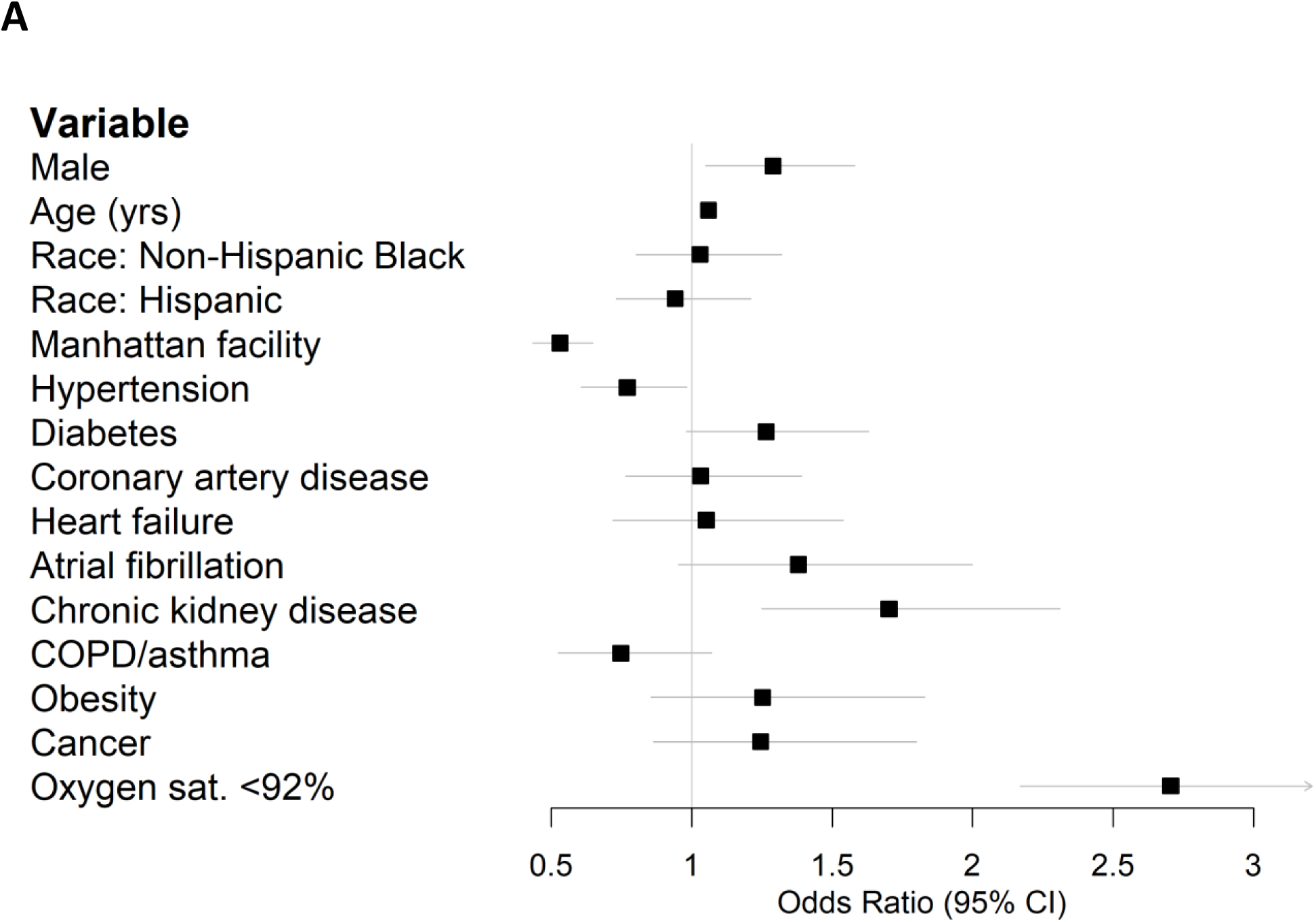
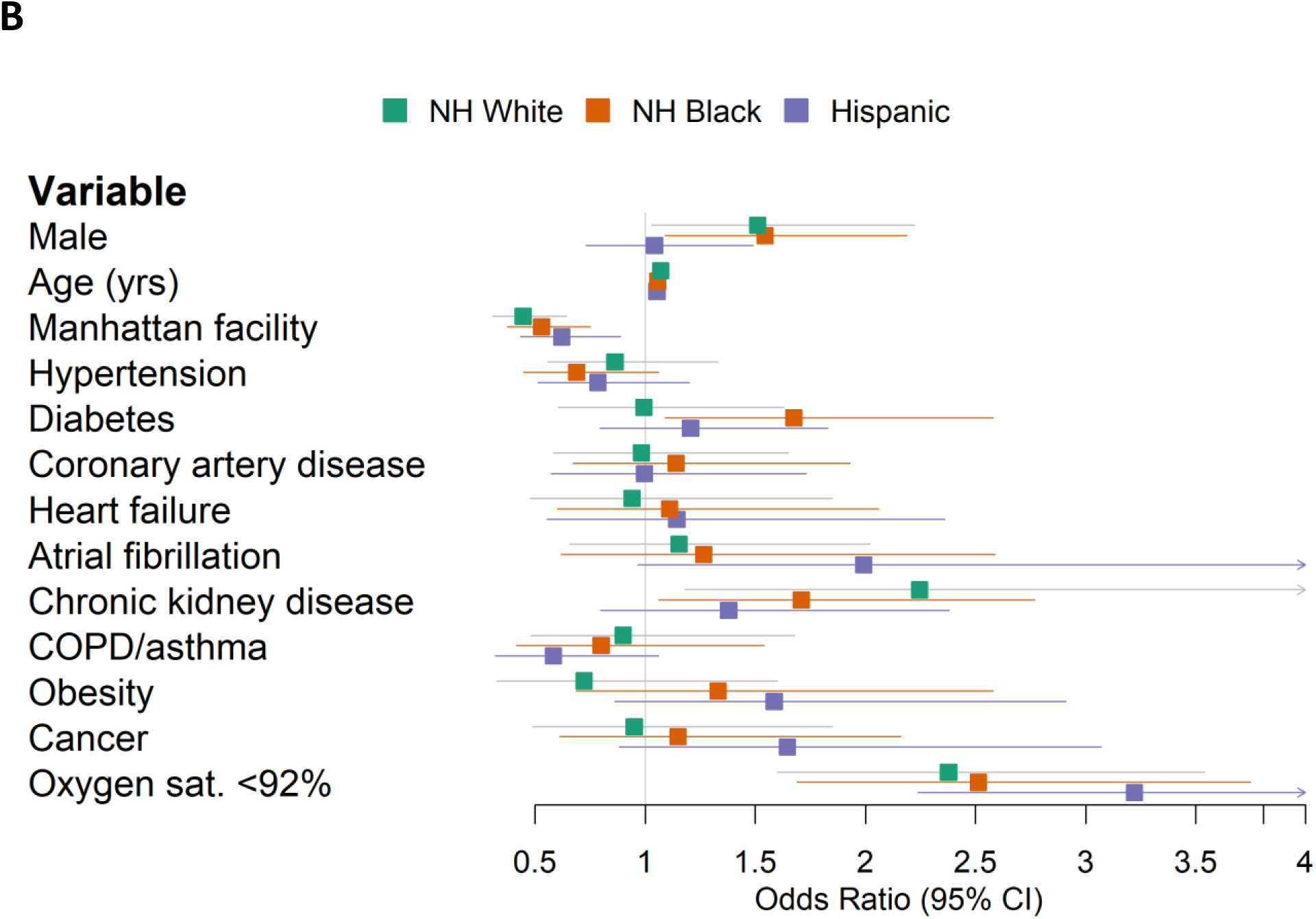
Forest plots of multivariable logistic regression results predicting in-hospital mortality. (A) All patients. (B) Stratified by race/ethnicity.

Previous patient cohort analyses rarely considered race/ethnic-specific risk factors, which require stratified modeling within each population cohort. We conducted stratified analyses within ethnic groups to determine the population-specific effect sizes of clinical factors and comorbidity. Diabetes was associated with an odds ratio of 1.67 (95% CI 1.09-2.58) in the stratified NH Black population, and an odds ratio of 0.99 (95% CI 0.61-1.63) in the NH White population (**Figure 2B**). Obesity was associated with an odds ratio of 1.33 (95% CI 0.69-2.58) in NH Black, compared to an odds ratio of 0.72 (95% CI 0.33-1.60) in NH White (**Figure 2B**). Increased age and baseline hypoxia were consistently associated with increased mortality across all three populations (**Supplemental Table 6**). Altogether, these results highlight the shared and specific clinical risk factors of COVID-19 mortality across populations.

### Baseline laboratory values

We analyzed baseline lab values among patients admitted to the largest site in our dataset, the Mount Sinai Hospital (MSH). This site had the most complete records for routine and inflammatory lab values. We defined baseline labs as the first lab value within 24 hours of the start of the encounter.

Among common lab values, Hispanic patients had higher baseline alanine aminotransferase values (ALT, 33 vs. 28 U/L, p=0.03) than NH White patients (Table 3), consistent with the increased prevalence of chronic liver disease among Hispanic patients.

**Table 3:** Baseline laboratory values among patients admitted to The Mount Sinai Hospital, by race/ethnicity

| Laboratory test | Evaluable Patients | NH White | NH Black | Hispanic |
| --- | --- | --- | --- | --- |
| White blood cells, $10^3/\mu\text{L}$ | 984 | 6.5 (5 - 9.825) | 6.3 (4.9 - 9.2) | 6.4 (5 - 9.1) |
| Hemoglobin, g/dL | 985 | 13.15 (12 - 14.125) | 12.6 (10.9 - 13.8)<sup>†</sup> | 13.2 (11.4 - 14.5) |
| Platelets, $10^3/\mu\text{L}$ | 980 | 205 (159.5 - 256) | 203 (157 - 264.25) | 212.5 (165.25 - 279.5) |
| Sodium, mmol/L | 984 | 136 (134 - 139) | 137 (135 - 140)<sup>†</sup> | 137 (134 - 139.25) |
| Potassium, mmol/L | 941 | 4 (3.7 - 4.55) | 4.3 (3.8 - 4.8)<sup>†</sup> | 4.2 (3.8 - 4.6) |
| Chloride, mmol/L | 984 | 101 (98 - 104) | 102 (97.25 - 105) | 102 (98 - 105) |
| Blood urea nitrogen, mg/dL | 984 | 17 (12 - 28) | 22 (14 - 46)<sup>†</sup> | 15 (10 - 27)<sup>†</sup> |
| Creatinine, mg/dL | 985 | 0.88 (0.7 - 1.22) | 1.25 (0.9 - 2.705)<sup>†</sup> | 0.87 (0.67 - 1.2325) |
| Aspartate aminotransferase, U/L | 915 | 38 (29 - 63) | 40 (28 - 59) | 43 (30 - 68.25) |
| Alanine aminotransferase, U/L | 958 | 28 (18 - 49) | 26 (16.5 - 41.5) | 33 (20 - 63)<sup>†</sup> |
| Total bilirubin, mg/dL | 966 | 0.6 (0.5 - 0.8) | 0.6 (0.4 - 0.9) | 0.6 (0.4 - 0.8) |
| Albumin, g/dL | 969 | 3.1 (2.7 - 3.5) | 3.2 (2.8 - 3.55) | 3.3 (2.9 - 3.6)† |
| D-dimer, ug/mL FEU | 755 | 1.25 (0.78 - 2.0475) | 1.59 (0.78 - 2.795) | 1.15 (0.6575 - 2.2125) |
| Ferritin, ng/mL | 904 | 679.5 (354.75 - 1423) | 717 (328 - 2124) | 665 (275.5 - 1649.75) |
| Ferritin, times upper limit of normal | 904 | 2.24 (1.190625 - 4.76625) | 3.16 (1.18 - 6.37)† | 2.66 (1.15 - 5.15) |
| Procalcitonin, ng/mL | 914 | 0.13 (0.06 - 0.375) | 0.29 (0.09 - 0.855)† | 0.17 (0.08 - 0.5075) |
| Lactate dehydrogenase, U/L | 854 | 405 (326 - 488) | 396.5 (312.5 - 574.75) | 388 (296 - 525) |
| C-reactive protein, mg/L | 903 | 119.35 (68.625 - 203.475) | 100.25 (49.45 - 173.45) | 100.4 (54.125 - 193.575) |
| Interleukin-1 beta, pg/mL | 418 | 0.5 (0.375 - 0.7) | 0.6 (0.4 - 0.9) | 0.5 (0.4 - 0.8) |
| Interleukin-6, pg/mL | 583 | 84.3 (44.8 - 149) | 69.05 (39.9 - 120.75) | 68.2 (40 - 135) |
| Interleukin-8, pg/mL | 521 | 41.25 (29.725 - 58.7) | 43.95 (24.725 - 70.15) | 43.85 (28.025 - 64) |
| Tumor necrosis factor-alpha, pg/mL | 522 | 24 (18.425 - 33) | 26.35 (17.6 - 45.4) | 22.3 (16.925 - 32.125) |

†: p<0.05 compared to White.

Among inflammatory lab markers, NH Black patients had higher initial levels of procalcitonin (0.29 vs. 0.13 ng/mL, p<0.001), and more abnormal ferritin levels (3.16 vs. 2.24 times upper limit of normal, p=0.03) compared to NH White patients (**Table 3**). There were no significant differences in baseline d-dimer, lactate dehydrogenase (LDH), C-reactive protein (CRP), interleukin-1-beta (IL-1B), interleukin-6 (IL-6), interleukin-8 (IL-8), or tumor necrosis factor-alpha (TNFa) levels. Thus, population-specific associations identified for these immune factors would indicate contributions from their relative differences between patients of different outcomes within a population rather than baseline differences across populations.

### Population-specific immune factors associated with clinical outcomes

We next conducted a multi-ethnic analysis to identify immune markers associated with patient outcomes in the MSH cohort. Using multivariate models adjusting for age, sex, and hypoxia, we identified CRP (OR 1.39, 95% CI 1.16-1.68), albumin (OR 0.75, 95% CI 0.61-0.91), IL-6 (OR 1.43, 95% CI 1.12-1.82), WBC (OR 1.35, 95% CI 1.08-1.65), and LDH (OR 1.34, 95% CI 1.07-1.68) as independent predictors of mortality (**Supplemental Table 7**).

To identify immune markers showing population-specificity in predicting COVID-19 outcomes, we applied the multivariate regression model in each population-stratified cohort. Elevated levels of IL-1B were associated with a higher risk of mortality in Black (OR 2.35, 95% CI 1.13-4.86) compared to White patients (OR 0.78, 95% CI 0.41-1.51) (**Figure 3, Supplemental Table 7**). Increased procalcitonin levels were associated with an odds ratio of 2.65 (95% CI 0.88-7.96) in Hispanic patients, compared to an odds ratio of 0.98 (95% CI 0.58-1.66) among NH White patients. Increased IL-8 levels were associated with an odds ratio of 1.51 (95% CI 0.59-3.86) among Hispanic patients, and an odds ratio of 8.76 (95% CI 0.95-80.7) among White patients (**Supplemental Table 7**). To validate the population-specificity of these COVID-19 mortality-associated immune markers, we further utilized a multivariate model including interaction terms, finding a significant interaction between NH Black and IL-1B (p=0.04) and a suggestive but non-significant interactions between Hispanic and procalcitonin (p=0.07) and IL-8 (p=0.09) compared to NH White (**Supplemental Table 6**).

**Figure 3:**
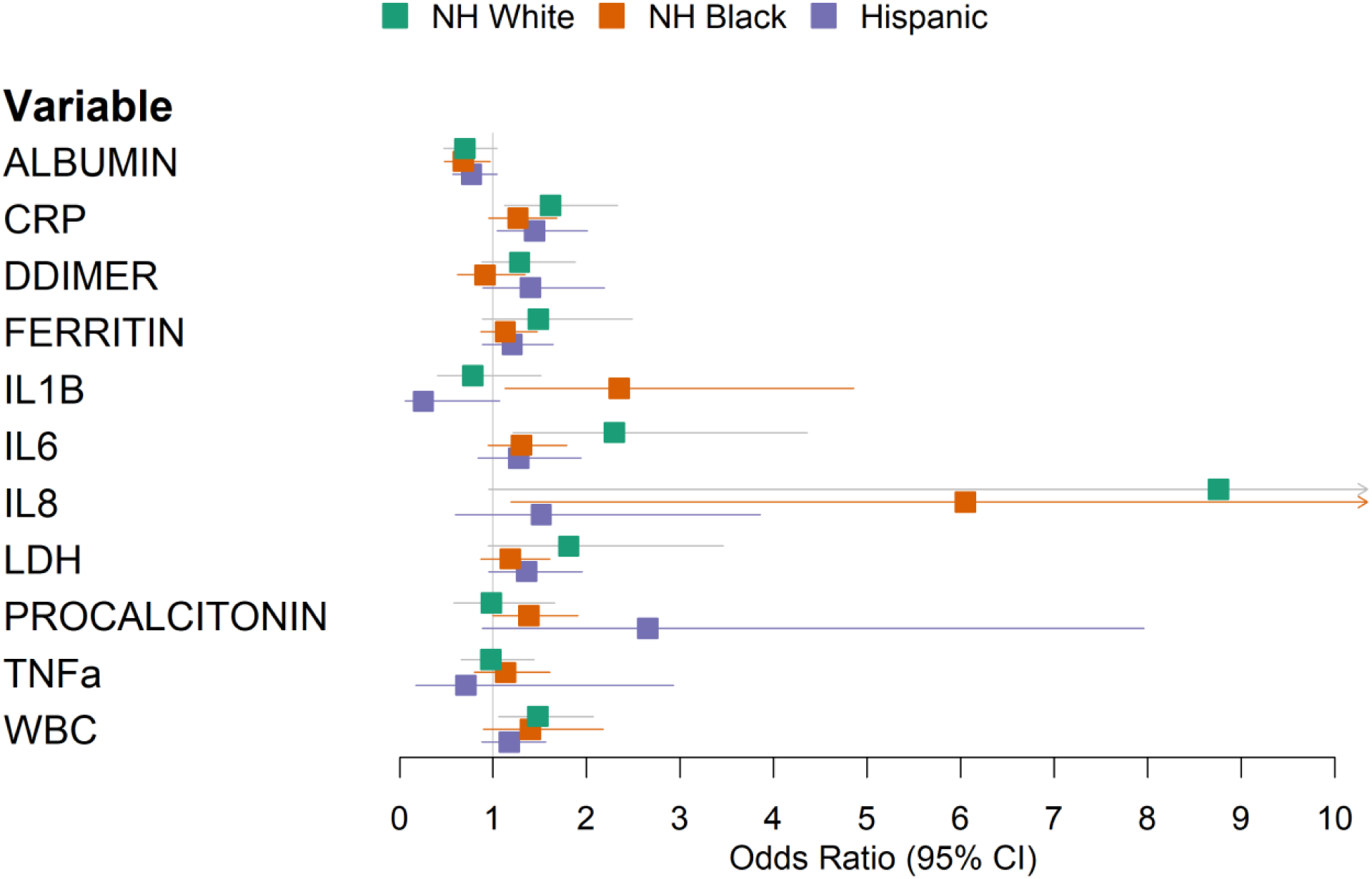
Forest plot of multivariable logistic regression results predicting in-hospital mortality using laboratory values, stratified by race/ethnicity. Models were adjusted for age, sex, and baseline hypoxia. NH: Non-Hispanic.

Next, we sought to validate the immune marker findings of the MSH cohort in an independent dataset. We utilized immunoprofiling data from the University of Pennsylvania cohort^15^ to compare levels of serum cytokines and immunologic markers between diverse patient populations vs. heatlhy and recovered donors. Among the COVID-19 patients with available race/ethnicity data, eight were NH Black, three were NH White, and four were Asian. Additionally, there were ten healthy donors (HDs) and twelve recovered donors (RDs) with no available race/ethnicity data.

We used the non-parametric Mood’s median test to detect potential population differences in the median values of 13 measured immune markers against the combined cohort of HDs and RDs (HDs/RDs, **Supplemental Table 8**). NH Black patients had significantly higher median IL-6 (37.1 vs 2.59, p=0.003) and IP10 (227 vs 55.6, p=0.006), and lower median IL12p70 (1.98 vs 3.41, p=0.004) levels than HDs/RDs. IP10 was also found to be significant when comparing NH White or Asian patients vs. HDs/RDs (p<0.05). Median IL-1B was 4.13 in NH Black patients compared to 2.66 in HDs/RDs (p=0.115), providing a suggestive yet non-significant association to the finding in the MSH cohort.

## Discussion

Racial disparities in COVID-19 infections and outcomes have become apparent in both the US and elsewhere.^1–4^ The causes of these disparities are complex and multifactorial and must be considered in the context of the social determinants of health.^4,16^

In this study, set in New York City during the height of the initial COVID-19 surge, we describe the characteristics and outcomes of a diverse cohort including substaintial numbers of NH White, NH Black, and Hispanic patients. The three groups differed significantly in demographic and clinical factors. White patients were older and showed higher rates of cardiovascular disease such as coronary artery disease atrial fibrillation. NH Black and Hispanic patients were younger, and had different comorbidity profiles, e.g. hypertension, diabetes, chronic kidney disease, and chronic liver disease.

Unadjusted in-hospital mortality was highest in NH White patients, but multivariable analysis showed that race/ethnicity was not an independent predictor of mortality in this cohort. It remains unclear whether race and ethnicity are independent risk factors for COVID-19 mortality after adjusting for confounding factors. Large national-level studies in the UK and Brazil have reported race as an independent predictor of mortality,^3,4^ whereas smaller studies in the US have not,^1,13,14^ possibly due to statistical power or population differences. Changes in clinical management and outcomes of COVID-19 over the course of the pandemic may also complicate comparisons of results from different time periods.^17^ In this New York City patient population, race/ethnicity was not an independent predictor for mortality.

In addition to describing this cohort, we aimed to test established COVID-19 risk factors for race/ethnicity-specific effects. Despite recapitulating several known risk factors, such as age, male sex, and hypoxia, we found only suggestive but non-significant interactions between Black race, diabetes and obesity, both of which tended to increase the mortality risk of Black patients to a greater degree than White patients. Notably, when analyzing inflammatory markers for their association with mortality, we found a significant interaction between the NH Black population and the inflammatory cytokine IL-1B.

Excessive inflammation has emerged as an important aspect of COVID-19 pathophysiology, and the anti-inflammatory steroid dexamethasone has been shown to improve outcomes among those with severe disease.^18^ The interaction between the NH Black population and IL-1B raises the possibility that differences in immunity may contribute to worse outcomes in some patients. Black Americans are at higher risk of autoimmune conditions such as systemic lupus erythematosus and lupus nephritis compared to White Americans, differences which can be linked in some cases to specific polymorphisms which are more common in African Americans.^19–22^

Interleukin-1-beta (IL-1B) is a pro-inflammatory cytokine that plays a role in both physiologic and pathologic inflammation. IL-1 inhibitors such as anakinra and canakinumab have been developed to target IL-1 in autoimmune diseases such as rheumatoid arthritis and Still’s disease. These agents are also being actively investigated as COVID-19 treatments. Thus far, two randomized studies have found no clinical benefit from IL-1 inhibitors in COVID-19.^23,24^ However, these studies have not reported analyses taking race/ethnicity into account.

The strengths of our database include its size and the inclusion of 37.1% Hispanic patients, a vulnerable population in this pandemic which, has been underrepresented in the literature to date. Additionally, our near-complete follow-up of the cohort’s hospital outcomes (99.3%) strengthens the validity of our findings.

In conclusion, our analysis of a diverse cohort drawn from the New York metropolitan area highlights both similarities and important differences across racial/ethnic groups in risk factors for death among hospitalized COVID-19 patients. The findings identified across populations call for conscious inclusion in future cohort studies and clinical trials to ensure the efficacy of potential diagnostics and treatments across diverse individuals.

## Limitations of the study

Our study has limitations that warrant specific mention. The cytokine analysis was limited to only a subset of the population and should be considered exploratory. We were not able to control for other comorbidities which may have influenced cytokine levels (e.g. diabetes and IL-1B).^25^ The dataset was derived from the electronic health record database without manual review, which may limit the completeness of comorbidity labels. Race and ethnicity were self-reported and were missing or unspecified in 17.4% of the initial cohort. The subset of patients with cytokine data was limited in number and models testing interactions Causal mechanisms underlying the correlations identified in this retrospective analysis remain to be elucidated.

## Methods

### Study setting

The study was conducted within the Mount Sinai Health System, which is an academic healthcare system comprising 8 hospitals and more than 410 ambulatory practice locations in the New York metropolitan area. This analysis involves patients who presented to five hospitals: The Mount Sinai Hospital (1,134 beds), Mount Sinai West (514 beds), and Mount Sinai Morningside (495 beds) in Manhattan; Mount Sinai Brooklyn (212 beds); and Mount Sinai Queens (235 beds).

### Data sources

Data were captured by the Epic electronic health record (Epic Systems, Verona, WI), and directly extracted from Epic’s Clarity and Caboodle servers. This de-identified dataset was developed and released by the Mount Sinai Data Warehouse (MSDW) team, with the goal of encompassing all COVID-19 related patient encounters within the Mount Sinai system, accompanied by selected demographics, comorbidities, vital signs, medications, and lab values. As part of de-identification, all patients over the age of 89 had their age set to 90.

This study utilized de-identified data extracted from the electronic health record and as such was considered non-human subject research. Therefore, this study was granted an exemption from the Mount Sinai IRB review and approval process.

### Patient population and definitions

The MSDW dataset captured any patient encounters at a Mount Sinai facility with any of the following: a COVID-19 related encounter diagnosis, a COVID-19 related visit type, a SARS-CoV-2 lab order, a SARS-CoV-2 lab result, or a SARS-CoV-2 lab test result from the New York State Department of Health’s Wadsworth laboratory. For this study, we identified patients with COVID-19 related visits to to the emergency department on or before April 13^th^, 2020 and selected patients who were admitted. We followed their hospitalization outcomes through June 2^nd^, 2020.

Our analysis was limited to adults over 18 years old who were hospitalized for COVID-19 through a Mount Sinai emergency department. Self-reported race and ethnicity were classified into 3 mutually exclusive categories: Non-Hispanic White (White), Non-Hispanic Black (Black), and Hispanic (**Supplemental Table 1**). COVID-19 positivity was determined by a positive or presumptive positive result from a nucleic acid-based test for SARS-CoV-2 in nasopharyngeal or oropharyngeal swab specimens. Baseline vital signs were the first documented vital signs for the encounter. Hypoxia was defined as an oxygen saturation less than 92%. We defined baseline labs as the first lab value within 24 hours of the start of the encounter.

### University of Pennsylvania cohort

Patients in the University of Pennsylvania cohort were identified based on a postive SARS-CoV-2 PCR test. Patients were screened and consented within 72 hours of hospitalization. Clinical data were collected from electronic medical records into standardized case reports. Healthy donors had no prior diagnosis or symptoms consident with COVID-19. Recovered donors were adults with a self reported positive COVID-19 PCR test who recovered as defined by the Centers for Disease Control. Cyotkine levels were measured from peripheral blood plasma using a custom human cytokine 31-plex panel (EMD Millipore Corporation, SPRCUS707), as described in Divij et al.^15^

### Logistic regression

The primary outcome was in-hospital mortality. Univariable and multivariable logistic regression were used to identify factors associated with death. Race/ethnicity-specific risk factors were identified by 1) constructing stratified models for each racial category, and 2) constructing models including interaction terms between race/ethnicity and other covariates. Separate interaction models were created to test the interactions of either Hispanic ethnicity or Black race with other covariates. Interactions were compared against White race as the reference group.

We analyzed demographic factors, comorbidities, initial vital signs, baseline lab values, and treatment facility site (Manhattan vs. Brooklyn/Queens) as covariates. There was minimal clustering of outcomes by treatment site (ICC (ρ) = 0.026), and this was modeled as a fixed effect. Covariates were chosen a *priori* based on prior reports. We report the odds ratios derived from the coefficients of each model, along with the Wald-type confidence interval, and p-values.

### Laboratory value analysis

Markers of inflammation, such as C-reactive protein (CRP), ferritin, and d-dimer, have been proposed as being correlated with COVID-19 severity. However, the missingness of these lab values varied across sites. Given the possibility of confounding by indication (if providers ordered these labs in more acutely ill patients), we limited the analyses involving lab tests to those obtained at the largest site (The Mount Sinai Hospital) and which had less than 15% missing values at that site. The cytokines interleukin-1-beta, interleukin-6, interleukin-8, and tumor necrosis factor-alpha were exempted from this threshold because they were obtained on a subset of COVID-19 patients in the context of a study with broad inclusion criteria.^26,27^

To test the associations of these lab values with mortality, we tested each lab test in race/ethnicity-stratified multivariable logistic regression models adjusting for age, sex, and hypoxia. The number of covariates in the model was limited due to the reduced sample size. Labs were standardized to a mean of 0 and a standard deviation of 1 prior to regression analysis.

### Statistical analysis

Patient characteristics and baseline vitals and labs were described using medians and ranges for continuous variables, and proportions for categorical variables. Continuous variables were compared using the Wilcoxon rank-sum test, and categorical variables were compared using Fisher’s exact test. All statistical analyses and data visualizations were carried out using R 4.0.0 (The R foundation, Vienna, Austria), along with the *tidyverse, ggpubr, forestplot*, and *Hmisc* packages. Statistical significance was defined as p<0.05.

### Patient and public involvement

Patients and the public were not involved in the design or conduct of this study.

## Data Availability

All data produced in the present study are available upon reasonable request to the authors

## Acknowledgment

We dedicate this work to the frontline health workers and staff of the Mount Sinai Healthcare System.

## Supplemental Materials

*Supplemental Table 1: Baseline demographic and clinical characteristics of patients presenting to the emergency department, by hospitalization status.*

*Supplemental Table 2: Baseline demographic and clinical characteristics of patients not admitted to the hospital, by race/ethnicity.*

*Supplemental Table 3: Self-reported ethnicities which were classified as (A) Hispanic, and (B) Non-Hispanic Black.*

*Supplemental Table 4: Univariable logistic regression using demographic and clinical factors to predict in-hospital mortality, stratified by race/ethnicity.*

*Supplemental Table 5: Univariable logistic regression using standardized laboratory values to predict in-hospital mortality, stratified by race/ethnicity.*

*Supplemental Table 6: Interaction p-values from multivariable logistic regression including interaction terms between race/ethnicity and demographic, clinical, or laboratory factors.*

*Supplemental Table 7: Multivariable logistic regression using standardized laboratory values to predict in-hospital mortality, stratified by race/ethnicity, adjusting for age, sex, and baseline hypoxia.*

*Supplemental Table 8: Median cytokine levels of patients from the University of Pennsylvania cohort.*

