## Supplemental Table 4 for "Multi-ethnic Investigation of Risk and Immune Determinants of COVID-19 Outcomes"

*Supplemental Table 4: Univariable logistic regression using demographic and clinical factors to predict in-hospital mortality, stratified by race/ethnicity.*

| Variable | White OR (95% CI) | Black OR (95% CI) | Hispanic OR (95% CI) | All patients OR (95% CI) |
|---|---|---|---|---|
| **Age (yrs)** | 1.07 (1.05-1.08) | 1.05 (1.04-1.07) | 1.05 (1.04-1.06) | 1.06 (1.05-1.06) |
| **Age 55-64** | 1.66 (0.715-3.85) | 2.51 (1.27-4.99) | 2.77 (1.65-4.66) | 2.38 (1.65-3.43) |
| **Age 65-74** | 3.56 (1.75-7.19) | 5 (2.61-9.62) | 4.22 (2.58-6.97) | 4.22 (3.03-5.94) |
| **Age >=75** | 10.3 (5.33-20) | 7.92 (4.16-15.1) | 6.05 (3.75-9.69) | 8.17 (5.91-11.2) |
| **Race: Non-Hispanic Black** | NA | NA | NA | 0.733 (0.589-0.913) |
| **Race: Hispanic** | NA | NA | NA | 0.606 (0.486-0.755) |
| **Manhattan facility** | 0.378 (0.274-0.523) | 0.517 (0.379-0.705) | 0.593 (0.431-0.816) | 0.477 (0.397-0.571) |
| **Current or former smoker** | 1.01 (0.702-1.46) | 0.933 (0.662-1.31) | 1.46 (1.03-2.06) | 1.11 (0.907-1.36) |
| **Hypertension** | 1.39 (0.998-1.94) | 1.1 (0.807-1.5) | 1.6 (1.17-2.2) | 1.32 (1.1-1.58) |
| **Diabetes** | 1.2 (0.795-1.8) | 1.57 (1.13-2.19) | 1.51 (1.08-2.1) | 1.37 (1.12-1.67) |
| **Coronary artery disease** | 1.6 (1.07-2.4) | 1.79 (1.16-2.77) | 1.65 (1.05-2.58) | 1.73 (1.36-2.22) |
| **Heart failure** | 1.85 (1.08-3.16) | 1.39 (0.826-2.33) | 1.52 (0.864-2.69) | 1.59 (1.17-2.17) |
| **Atrial fibrillation** | 1.86 (1.18-2.95) | 1.94 (1.02-3.68) | 3.06 (1.62-5.83) | 2.31 (1.68-3.18) |
| **Chronic kidney disease** | 2.32 (1.33-4.04) | 1.75 (1.19-2.59) | 1.45 (0.944-2.23) | 1.65 (1.28-2.12) |
| **COPD/asthma** | 1.13 (0.652-1.96) | 0.634 (0.352-1.14) | 0.925 (0.541-1.58) | 0.87 (0.634-1.2) |
| **Obesity** | 0.523 (0.262-1.04) | 0.817 (0.47-1.42) | 1.32 (0.793-2.2) | 0.861 (0.62-1.2) |
| **Cancer** | 1.02 (0.566-1.86) | 1.15 (0.656-2.02) | 1.95 (1.11-3.43) | 1.33 (0.959-1.85) |
| **Respiratory rate** | 1.03 (1.01-1.06) | 1.1 (1.07-1.14) | 1.1 (1.07-1.13) | 1.08 (1.06-1.09) |
| **Oxygen sat. <92%** | 2.09 (1.48-2.96) | 2.72 (1.89-3.94) | 2.71 (1.96-3.74) | 2.43 (2-2.95) |
