## Supplemental Table 5 for "Multi-ethnic Investigation of Risk and Immune Determinants of COVID-19 Outcomes"

*Supplemental Table 5: Univariable logistic regression using standardized laboratory values to predict in-hospital mortality, stratified by race/ethnicity.*

| Test | White OR (95% CI) | Black OR (95% CI) | Hispanic OR (95% CI) | All patients OR (95% CI) |
|---|---|---|---|---|
| **WBC** | 1.62 (1.17-2.24) | 1.53 (1.04-2.25) | 1.32 (0.994-1.75) | 1.48 (1.22-1.79) |
| **Albumin** | 0.589 (0.408-0.85) | 0.587 (0.425-0.81) | 0.673 (0.513-0.883) | 0.624 (0.522-0.747) |
| **D-dimer** | 1.37 (0.971-1.94) | 1.17 (0.854-1.59) | 1.79 (1.17-2.74) | 1.33 (1.1-1.62) |
| **Ferritin** | 1.36 (0.821-2.24) | 1.1 (0.858-1.4) | 1.18 (0.916-1.51) | 1.15 (0.977-1.35) |
| **Procalcitonin** | 1.01 (0.644-1.59) | 1.37 (1.01-1.85) | 2.71 (0.965-7.63) | 1.31 (1.07-1.61) |
| **LDH** | 1.72 (0.951-3.12) | 1.3 (0.97-1.73) | 1.45 (1.04-2.01) | 1.38 (1.13-1.7) |
| **CRP** | 1.65 (1.18-2.3) | 1.31 (0.995-1.72) | 1.66 (1.24-2.22) | 1.52 (1.28-1.81) |
| **IL-1B** | 0.879 (0.515-1.5) | 2.06 (1.09-3.87) | 0.411 (0.121-1.4) | 1.01 (0.774-1.32) |
| **IL-6** | 2.55 (1.36-4.76) | 1.39 (1.03-1.87) | 1.48 (1-2.18) | 1.57 (1.23-2) |
| **IL-8** | 12.9 (1.72-97.9) | 7.69 (1.62-36.2) | 2.18 (0.792-6.01) | 4.31 (1.76-10.5) |
| **TNF-alpha** | 0.999 (0.741-1.35) | 1.25 (0.893-1.75) | 0.987 (0.314-3.1) | 1.09 (0.918-1.3) |
