## Supplemental Table 6 for "Multi-ethnic Investigation of Risk and Immune Determinants of COVID-19 Outcomes"

*Supplemental Table 6: Interaction p-values from multivariable logistic regression including interaction terms between race/ethnicity and demographic, clinical, or laboratory factors.*

| Variable | Black vs. White Interaction p-value | Hispanic vs. White Interaction p-value |
|---|---|---|
| Age (yrs) | 0.157 | 0.153 |
| Manhattan facility | 0.477 | 0.176 |
| Hypertension | 0.938 | 0.964 |
| Diabetes | 0.0885 | 0.629 |
| Coronary artery disease | 0.597 | 0.833 |
| Heart failure | 0.535 | 0.853 |
| Atrial fibrillation | 0.786 | 0.347 |
| Chronic kidney disease | 0.783 | 0.206 |
| Obesity | 0.1 | 0.0647 |
| Cancer | 0.67 | 0.266 |
| Oxygen sat. <92% | 0.831 | 0.192 |
| **Laboratory Values** | | |
| Albumin | 0.72 | 0.817 |
| CRP | 0.473 | 0.917 |
| D-dimer | 0.319 | 0.707 |
| Ferritin | 0.324 | 0.441 |
| IL-1B | 0.038 | 0.167 |
| IL-6 | 0.158 | 0.19 |
| IL-8 | 0.752 | 0.0862 |
| LDH | 0.266 | 0.585 |
| Procalcitonin | 0.22 | 0.0734 |
| TNF-alpha | 0.573 | 0.544 |
| WBC | 0.954 | 0.471 |
