## Supplemental Table 7 for "Multi-ethnic Investigation of Risk and Immune Determinants of COVID-19 Outcomes"

*Supplemental Table 7: Multivariable logistic regression using standardized laboratory values to predict in-hospital mortality, stratified by race/ethnicity, adjusting for age, sex, and baseline hypoxia.*

| var_label | White OR (95% CI) | Black OR (95% CI) | Hispanic OR (95% CI) | All patients OR (95% CI) |
|---|---|---|---|---|
| **Albumin** | 0.698 (0.47-1.04) | 0.681 (0.479-0.968) | 0.769 (0.568-1.04) | 0.745 (0.612-0.906) |
| **CRP** | 1.613 (1.12-2.33) | 1.266 (0.954-1.68) | 1.445 (1.04-2.01) | 1.392 (1.16-1.68) |
| **D-dimer** | 1.283 (0.878-1.88) | 0.911 (0.62-1.34) | 1.398 (0.892-2.19) | 1.141 (0.917-1.42) |
| **Ferritin** | 1.484 (0.886-2.49) | 1.13 (0.866-1.47) | 1.204 (0.885-1.64) | 1.186 (0.991-1.42) |
| **IL-1B** | 0.782 (0.405-1.51) | 2.349 (1.13-4.86) | 0.254 (0.0596-1.07) | 0.977 (0.741-1.29) |
| **IL-6** | 2.296 (1.21-4.36) | 1.302 (0.949-1.79) | 1.274 (0.837-1.94) | 1.43 (1.12-1.82) |
| **IL-8** | 8.758 (0.953-80.7) | 6.05 (1.19-30.3) | 1.514 (0.594-3.86) | 2.858 (1.14-7.17) |
| **LDH** | 1.808 (0.945-3.46) | 1.183 (0.866-1.61) | 1.359 (0.95-1.95) | 1.344 (1.07-1.68) |
| **Procalcitonin** | 0.981 (0.581-1.66) | 1.383 (0.999-1.91) | 2.654 (0.884-7.96) | 1.27 (1.01-1.59) |
| **TNF-alpha** | 0.974 (0.66-1.44) | 1.132 (0.798-1.61) | 0.71 (0.172-2.93) | 1.052 (0.863-1.28) |
| **WBC** | 1.478 (1.06-2.07) | 1.399 (0.898-2.18) | 1.172 (0.879-1.56) | 1.335 (1.08-1.65) |
