## Supplemental Table 8 for "Multi-ethnic Investigation of Risk and Immune Determinants of COVID-19 Outcomes"

*Supplemental Table 8: Median cytokine levels of patients from the University of Pennsylvania cohort.*

| Cytokine | Healthy Donor (HD, N=10) | Recovered Donor (RD, N=12) | HD+RD (N=22) | NH Black (NHB, N=8) | Asian (AS, N=4) | NH White (NHW, N=3) | P-value (NHB vs. HD+RD) | P-value (NHW vs. HD+RD) | P-value (AS vs. HD+RD) |
|---|---|---|---|---|---|---|---|---|---|
| IL1beta | 2.20 | 3.88 | 2.66 | 4.13 | 4.44 | 3.8 | 0.115 | 0.138 | 0.090 |
| IL6 | 2.25 | 2.59 | 2.53 | 37.1 | 54.6 | 211 | **0.003** | 0.157 | 0.094 |
| TNFa | 14.2 | 12.6 | 13.7 | 9.47 | 7.37 | 9.15 | 0.165 | 0.086 | 0.231 |
| IP10 | 77.2 | 55.6 | 61.5 | 227 | 266 | 509 | **0.006** | **0.041** | **0.025** |
| IFNlamda1 | 82.5 | 82.1 | 82.1 | 71.1 | 90.7 | 49.0 | 0.659 | 0.863 | 0.514 |
| IL8 | 8.25 | 6.74 | 7.60 | 18.5 | 12.8 | 36.2 | 0.659 | 0.157 | 0.094 |
| IL12p70 | 3.14 | 3.42 | 3.20 | 1.98 | 1.78 | 4.57 | **0.004** | 0.707 | 0.513 |
| IFNa2 | 2.72 | 4.82 | 3.49 | 5.06 | 4.92 | 3.90 | 0.638 | 0.707 | 0.497 |
| IFNlamda2.3 | 12.2 | 15.2 | 13.7 | 12.3 | 12.0 | 12.9 | 0.693 | 0.598 | 0.693 |
| GMCSF | 11.7 | 7.15 | 8.32 | 6.57 | 7.92 | 8.44 | 0.296 | 0.598 | 0.693 |
| IFNbeta | 4.25 | 4.25 | 4.25 | 7.07 | 5.39 | 4.25 | 0.650 | 0.985 | 0.260 |
| IL10 | 12.3 | 9.34 | 10.5 | 15.5 | 18.8 | 18.3 | 0.189 | 0.117 | 0.069 |
| IFNg | 5.97 | 7.91 | 6.04 | 7.32 | 8.71 | 7.36 | 0.296 | 0.328 | 0.231 |
