## Supplemental Table 1 for "Multi-ethnic Investigation of Risk and Immune Determinants of COVID-19 Outcomes"

Supplemental Table 1: Baseline demographic and clinical characteristics of patients presenting to the emergency department, by hospitalization status.

| | Admitted (N=3,086) | Not Admitted (N=1,911) | P-value |
|---|---|---|---|
| **Age (yrs)** | 66 (56 - 77) | 50 (36 - 62) | <0.001 |
| **Asian** | 144 (4.8%) | 86 (4.6%) | 0.781 |
| **Hispanic** | 892 (29.7%) | 495 (26.4%) | 0.014 |
| **Non-Hispanic Black** | 825 (27.4%) | 635 (33.9%) | <0.001 |
| **Non-Hispanic White** | 689 (22.9%) | 397 (21.2%) | 0.168 |
| **Other** | 458 (15.2%) | 262 (14%) | 0.245 |
| **Current smoker** | 113 (4.7%) | 78 (5.5%) | 0.282 |
| **Former smoker** | 658 (27.4%) | 204 (14.4%) | <0.001 |
| **Never smoker** | 1629 (67.9%) | 1133 (80.1%) | <0.001 |
| **Hypertension** | 1096 (35.5%) | 258 (13.5%) | <0.001 |
| **Diabetes** | 741 (24%) | 149 (7.8%) | <0.001 |
| **Coronary artery disease** | 395 (12.8%) | 78 (4.1%) | <0.001 |
| **Heart failure** | 218 (7.1%) | 36 (1.9%) | <0.001 |
| **Atrial fibrillation** | 201 (6.5%) | 36 (1.9%) | <0.001 |
| **Chronic kidney disease** | 368 (11.9%) | 59 (3.1%) | <0.001 |
| **COPD/asthma** | 265 (8.6%) | 86 (4.5%) | <0.001 |
| **Obesity** | 250 (8.1%) | 91 (4.8%) | <0.001 |
| **Cancer** | 205 (6.6%) | 59 (3.1%) | <0.001 |
| **Chronic liver disease** | 83 (2.7%) | 28 (1.5%) | 0.004 |
| **Obstructive sleep apnea** | 65 (2.1%) | 19 (1%) | 0.003 |
| **HIV** | 56 (1.8%) | 21 (1.1%) | 0.058 |
| **Temperature (°F)** | 98.9 (98.2 - 100.2) | 98.6 (97.9 - 99.7) | <0.001 |
| **Heart rate (bpm)** | 97 (84 - 110) | 92 (81 - 104) | <0.001 |
| **Systolic blood pressure (mmHg)** | 129 (115 - 146) | 131 (119 - 144) | 0.02 |
| **Respiratory rate (bpm)** | 20 (18 - 22) | 18 (17 - 20) | <0.001 |
| **Oxygen saturation (%)** | 95 (91 - 97) | 98 (96 - 99) | <0.001 |
| **Oxygen sat. <92%** | 775 (25.1%) | 96 (5%) | <0.001 |
