## Supplemental Table 2 for "Multi-ethnic Investigation of Risk and Immune Determinants of COVID-19 Outcomes"

Supplemental Table 2: Baseline demographic and clinical characteristics of patients presenting to the emergency department but not admitted to the hospital, by race/ethnicity.

| | NH Black (N=635) | NH White (N=397) | Hispanic (N=495)) | Asian (N=86) | Other (N=262) |
|---|---|---|---|---|---|
| **Age (yrs)** | 50 (37 - 61) | 52 (35 - 64) | 52 (35 - 63) | 47.5 (34.25 - 60) | 49 (36 - 61) |
| **Current smoker** | 38 (7.5%) | 12 (4.3%) | 18 (5.2%) | 4 (6.2%) | 5 (2.5%) |
| **Former smoker** | 55 (10.9%) | 46 (16.4%) | 68 (19.7%) | 5 (7.8%) | 29 (14.6%) |
| **Never smoker** | 412 (81.6%) | 223 (79.4%) | 259 (75.1%) | 55 (85.9%) | 164 (82.8%) |
| **Hypertension** | 90 (14.2%) | 43 (10.8%) | 87 (17.6%) | 12 (14%) | 24 (9.2%) |
| **Diabetes** | 49 (7.7%) | 21 (5.3%) | 48 (9.7%) | 8 (9.3%) | 22 (8.4%) |
| **Coronary artery disease** | 19 (3%) | 22 (5.5%) | 25 (5.1%) | 1 (1.2%) | 11 (4.2%) |
| **Heart failure** | 10 (1.6%) | 7 (1.8%) | 15 (3%) | 1 (1.2%) | 3 (1.1%) |
| **Atrial fibrillation** | 6 (0.9%) | 13 (3.3%) | 9 (1.8%) | NA | 8 (3.1%) |
| **Chronic kidney disease** | 22 (3.5%) | 9 (2.3%) | 20 (4%) | NA | 8 (3.1%) |
| **COPD/asthma** | 34 (5.4%) | 10 (2.5%) | 33 (6.7%) | NA | 8 (3.1%) |
| **Obesity** | 37 (5.8%) | 5 (1.3%) | 37 (7.5%) | 2 (2.3%) | 10 (3.8%) |
| **Cancer** | 11 (1.7%) | 13 (3.3%) | 25 (5.1%) | 1 (1.2%) | 9 (3.4%) |
| **Chronic liver disease** | 8 (1.3%) | 6 (1.5%) | 7 (1.4%) | 1 (1.2%) | 4 (1.5%) |
| **Obstructive sleep apnea** | 10 (1.6%) | 1 (0.3%) | 4 (0.8%) | NA | 4 (1.5%) |
| **HIV** | 5 (0.8%) | 2 (0.5%) | 12 (2.4%) | NA | 2 (0.8%) |
| **Temperature (°F)** | 98.5 (97.8 - 99.6) | 98.25 (97.7 - 99.1) | 99 (98.3 - 100.4) | 98.6 (97.9 - 99.5) | 98.7 (98 - 100) |
| **Heart rate (bpm)** | 92 (82 - 102) | 88 (78 - 99) | 95 (82.5 - 106) | 94 (81 - 105.75) | 96 (82.25 - 108) |
| **Systolic blood pressure (mmHg)** | 133 (120.5 - 148) | 130 (118 - 141) | 130 (119 - 144) | 129 (118 - 143.5) | 129.5 (117 - 140.75) |
| **Respiratory rate (bpm)** | 18 (17 - 19) | 18 (17 - 20) | 18 (18 - 20) | 18 (16 - 19) | 18 (18 - 20) |
| **Oxygen saturation (%)** | 98 (97 - 100) | 98 (96 - 99) | 97 (96 - 98) | 98 (97 - 99) | 98 (96 - 99) |
| **Oxygen sat. <92%** | 24 (3.8%) | 16 (4%) | 32 (6.5%) | 6 (7.1%) | 16 (6.1%) |
