## Supplemental Table 3 for "Multi-ethnic Investigation of Risk and Immune Determinants of COVID-19 Outcomes"

Supplemental Table 3: Self-reported ethnicities which were classified as (A) Hispanic, and (B) Non-Hispanic Black.

(A) Self-reported ethnicities appearing in the dataset which were classified as Hispanic:

- ARGENTINEAN
- BOLIVIAN
- CASTILLIAN
- CENTRAL AMERICAN
- CHICANO
- CHILEAN
- COLOMBIAN
- COSTA RICAN
- CUBAN
- DOMINICAN
- ECUADORIAN
- GUATEMALAN
- HONDURAN
- LATIN AMERICAN
- MEXICAN
- MEXICAN AMERICAN
- MEXICANO
- NICARAGUAN
- PANAMANIAN
- PARAGUAYAN
- PERUVIAN
- PUERTO RICAN
- SALVADORAN
- SOUTH AMERICAN
- SPANIARD
- SPANISH BASQUE
- VENEZUELAN

(B) Self-reported races appearing in the dataset which were classified as Non-Hispanic Black (if lacking a Hispanic ethnicity):

- BARBADIAN
- CAPE VERDIAN
- CONGOLESE
- DOMINICA ISLANDER
- ERITREAN
- ETHIOPIAN
- GABONIAN
- GHANAIAN
- GRENADIAN
- GUINEAN
- HAITIAN
- IVORY COASTIAN
- JAMAICAN
- KENYAN
- LIBERIAN
- MADAGASCAR
- MALIAN
- NIGERIAN
- OTHER: EAST AFRICAN
- OTHER: NORTH AFRICAN
- OTHER: SOUTH AFRICAN
- OTHER: WEST AFRICAN
- SENEGALESE
- SIERRA LEONEAN
- SOMALIAN
- ST VINCENTIAN
- SUDANESE
- TANZANIAN
- TRINIDADIAN
- UGANDAN
- WEST INDIAN
- ZIMBABWEAN
